# Anthropometric-related percentile curves for muscle size and strength of lower limb muscles of typically developing children

**DOI:** 10.1101/2024.03.27.24304866

**Authors:** Ines Vandekerckhove, Britta Hanssen, Nicky Peeters, Tijl Dewit, Nathalie De Beukelaer, Marleen Van den Hauwe, Liesbeth De Waele, Anja Van Campenhout, Friedl De Groote, Kaat Desloovere

**Author notes:** **Corresponding author**: Kaat Desloovere, Weligerveld 1, 3212 Pellenberg (Belgium). These authors share first authorship (Ines Vandekerckhove, Britta Hanssen). Part of the material within this paper has been presented as a conference abstract at the ESMAC annual conference 2023 in Athens (21^st^ to 23^rd^ of September).

## Abstract

**Aims:** To establish anthropometric-related percentile curves for muscle size and strength in a cohort of typically developing (TD) children and to demonstrate their use through applications in children with cerebral palsy (CP) and Duchenne muscular dystrophy (DMD).

**Methods:** Lower limb muscle size and strength were assessed in a large cross-sectional cohort of TD children with 3D freehand ultrasound (four muscles, n=154, male/female=82/72, age range: 0y7mo-17y10mo) and fixed dynamometry (seven muscle groups, n=153, male/female=108/45, age range: 4y6mo-16y1mo), respectively. Generalized additive models for location, scale and shape were used to estimate anthropometric-related, i.e. body mass and height, TD percentile curves and to convert outcomes of individual patients with CP and DMD into z-scores.

**Results:** Muscle size and strength, as well as their inter-subject variation, increased with increasing anthropometric values. Individual patients exhibited negative z-scores, indicating muscle size and strength deficits in reference to TD peers.

**Interpretation:** The established anthropometric-related percentile curves for muscle size and strength in a cohort of TD children can successfully be used to express patient outcomes in reference to TD. This facilitates the interpretation of muscle size and weakness in children with a motor disability and allows for the evaluation of the disease progression and treatment impact during growth.

## Introduction

Muscle size and muscle strength increase gradually throughout childhood.^1,2^ However, this increase can be significantly altered in children with motor disorders like cerebral palsy (CP) ^3–6^ and Duchenne muscular dystrophy (DMD).^5,7^ Therefore, it is important to assess these parameters such that the progression of these disorders can be monitored and the effectiveness of treatment can be evaluated.

CP is an umbrella term for one of the most prevalent neurological disorders in children.^8^ It is estimated to affect 1-2 per 1,000 live births in high income countries.^9^ CP is caused by a non-progressive brain lesion in the developing fetal or infant brain, resulting in disorders of movement, posture and motor function.^8^ The initial brain injury in children with CP results in an altered neural drive to skeletal muscles and subsequent alterations in movement.^10,11^ Consequently, children with CP often present with muscle weakness and their skeletal muscle tissue shows a lack of growth throughout childhood.^3,4,12^

DMD is a progressive X-linked muscular disorder, affecting 1 in 3,500-6000 new-born boys.^7,13^ It is due to a genetic mutation in the *DMD* gene causing a deficiency of the muscle-stabilizing protein dystrophin.^7^ This deficiency leads to muscle degeneration, which is characterized by progressive loss of contractile tissue with replacement by fat and fibrotic tissue.^7,14^ This degeneration results in a progressive loss of muscle strength and alterations in posture and gait.^7,14^

Given the relationship between impaired muscle development and limitations in functioning and participation, close follow-up of muscular parameters is necessary. The quantification of these parameters is nowadays achievable through valid, reliable and clinically applicable techniques. Muscle strength can be evaluated with instrumented strength assessments, using dynamometry^5,15^, while muscle morphology can be evaluated with 3-dimensional freehand ultrasound.^16,17^

However, interpreting the impact of treatment or disease progression in growing children is challenging, as these effects are hard to distinguish from those of growth itself.^2^ Therefore, accounting for anthropometric growth is essential to enable a meaningful interpretation of individual outcomes. This can be done by matching a participant to a typically developing (TD) child. However, defining the parameters for matching and finding the appropriate match is a complex task, particularly because children with pediatric disorders often have altered anthropometric measures.^18,19^ Another approach is to normalize muscle parameters with respect to these anthropometric measures. Although this works well for certain age ranges and parameters, normalization is imperfect for wide age ranges and especially for very young ages.^1,4^

Growth percentile curves are widely used in child health and pediatric clinical practice to assess anthropometric measurements that vary by age.^20^ These charts can be utilized to express the level of pathology by converting assessed muscle size and strength outcomes into z-scores. Yet, so far, these TD percentile curves have not yet been established for these muscle outcomes. Therefore, the aim of the current study was to establish anthropometric-related percentile curves in a cohort of TD children covering childhood development, encompassing both muscle size and strength outcomes. The use of the TD percentile curves was demonstrated through application in children with CP and DMD.

## Methods

### Participants

A locally established cross-sectional reference database consisted of 154 TD children for muscle size (male/female=82/72, aged 0y7mo-17y10mo) and 153 TD children for muscle strength (male/female=108/45, aged 4y6mo-16y1mo). The recruitment occurred via schools in Belgium and via colleagues and students from the clinical motion analysis laboratory of the University Hospitals of Leuven (Pellenberg), KU Leuven and the University of Ghent. Children with a medical history of neurological, neuromuscular, or orthopedic disorders affecting the lower limbs or who performed organized sports at an intensive level (over 6 hours a week), were excluded.

The local ethics committees (Ethical Committee UZ Leuven/KU Leuven; s59945, s61324, s62187, s62645 and s63340 and Commissie voor Medische Ethiek-Universiteit Gent; EC/2017/0526) approved this study under the Declaration of Helsinki. Written informed consents were signed by participants’ parents/caregivers or participants of 18 years old. Participants above the age of 12 signed an informed assent.

### Data collection and analysis

Muscle size and strength were collected unilaterally and the assessed side was randomly selected.

### Anthropometric outcomes

The participants’ body mass, height and segment lengths of the lower limb were collected using standardized anthropometric methods.

### Muscle size

Muscle size of four lower limb muscles, m. rectus femoris, the distal part of the m. semitendinosus, m. tibialis anterior and m. medial gastrocnemius, was measured using 3D freehand ultrasound. The applied technique and protocol have previously been reported and were proven to be reliable.^17,21^ In brief, four reflective markers were attached on the linear probe of a Telemed-Echoblaster B-mode ultrasound device (Telemed-Echoblaster 128 Ext-1Z, with a 5.9-cm 10-MHz linear US transducer, Telemed Ltd., Vilnius, Lithuania) and were tracked with a motion tracking system (Optitrack V120:Trio, NaturalPoint Inc., Corvallis, Oregon, USA) to determine the orientation and position of the 2D ultrasound images. The Portico, i.e. a custom-made gel pad, was fixed on the ultrasound probe to minimize muscle deformation.^22^ STRADWIN software (version 6.0; Mechanical Engineering, Cambridge University, Cambridge, UK) was used for data collection and processing. Muscle volume (mL) was estimated by an automatic cubic planimetry interpolation of manually drawn transverse plane segmentations along the inside of the muscle border. Muscle belly length (mm) was calculated as Euclidean distance between relevant anatomical landmarks. Mid-belly anatomical cross-sectional area (mm^2^) was defined by a single segmentation at 50% of the muscle belly length. Ultrasound settings and parameter definition were applied as reported in earlier investigations.^1,17^

### Muscle strength

Muscle strength of seven lower limb muscle groups, namely the hip extensors, hip flexors, hip abductors, knee extensors, knee flexors, plantar flexors and dorsiflexors, was assessed according to a previously introduced, reliable and valid instrumented strength assessment.^5,15^ Maximal voluntary isometric contractions (MVIC) were performed using a fixed dynamometer (MicroFet, Hogan Health Industries, West Jordan, UT United States) in a custom-made chair. The maximal force (Newton [N]) per MVIC was extracted and averaged over one to three MVIC trials per muscle group. The mean maximal joint torque (Newton-meter [Nm]) for each muscle group was obtained by multiplying the mean maximal force with the lever arm determined at 75% of the segment length^15^. This analysis was performed with custom-written MATLAB (The Mathworks Inc., Natick, M.A., R2021b) scripts.

### Statistical Analysis

Anthropometric-related TD percentile curves for muscle size and strength were estimated using generalized additive models for location, scale and shape (GAMLSS), as suggested by the WHO Multicenter Growth Reference Study Group.^20,23,24^ GAMLSS allow modelling the parameters of the outcome distribution with respect to an explanatory variable. Similar anthropometric counterparts, which have often been used to normalize absolute outcomes ^25^, were chosen as explanatory variables to model muscle size and strength: body mass*height was selected for muscle volume and strength, height for muscle length, and body mass for cross-sectional area. Three different distributions were fitted to each outcome: (1) the Box–Cox Cole and Green distribution, or also known as the LMS method^26^, fits the skewness (lambda), median (mu) and coefficient of variation (sigma), while (2) the Box–Cox power exponential and (3) the Box–Cox t distributions, also known as the LMSP^27^ and LMST^28^ methods, respectively, enable fitting a fourth parameter, namely kurtosis (tau). Additionally, the effect of anthropometrics on the distribution parameters was modelled as a constant, as a linear function and as a P-spline. Model reduction was evaluated using the Schwartz Bayesian and the Akaike information criterions, while normalized quantile residual plots, summary statistics, worm plots and Q-statistics were used as diagnostic tools to select the final model including the fitted distribution with the modelled effect of anthropometrics on the distribution parameters.^23,24^ Moreover, the choice of the final model was also determined by checking whether the models were clinically realistic and biological plausible. Finally, the selected models were used to generate the curves for percentiles 5% (z-score=-1.645), 10% (z-score=-1.282), 25% (z-score=-0.675), 50% (z-score=0), 75% (z-score=0.675), 90% (z-score=1.282) and 95% (z-score=1.645) as well as to convert outcomes of individual patients into z-scores. All statistical analyses were performed using the GAMLSS package in R – 4.2.0 (R Core Team, Vienna, Austria). The TD percentile curves were digitized and an online graphical user interface (https://cmal.shinyapps.io/Z-score_calculator_MG_muscle_volume/) was developed to plot subject-specific muscle size and strength outcomes on the TD percentile curves and to convert these outcomes into z-scores. The platform is dynamic, as the TD percentile curves will be updated when more data becomes available.

## Results

Muscle volume and cross-sectional area increased stronger at small than large anthropometric values (except tibialis anterior cross-sectional area which increased linearly), while muscle belly length and strength increased linearly (except knee flexion and dorsiflexion strength which increased stronger at small than large anthropometric values) (Figures 1-3, Appendices S1-S2). The increase in inter-subject variation with anthropometric values was the largest for muscle volume and strength, then for cross-sectional area, and the least for than muscle belly length, whilst muscle strength showed already a high variation from the start.

**Figure 1:**
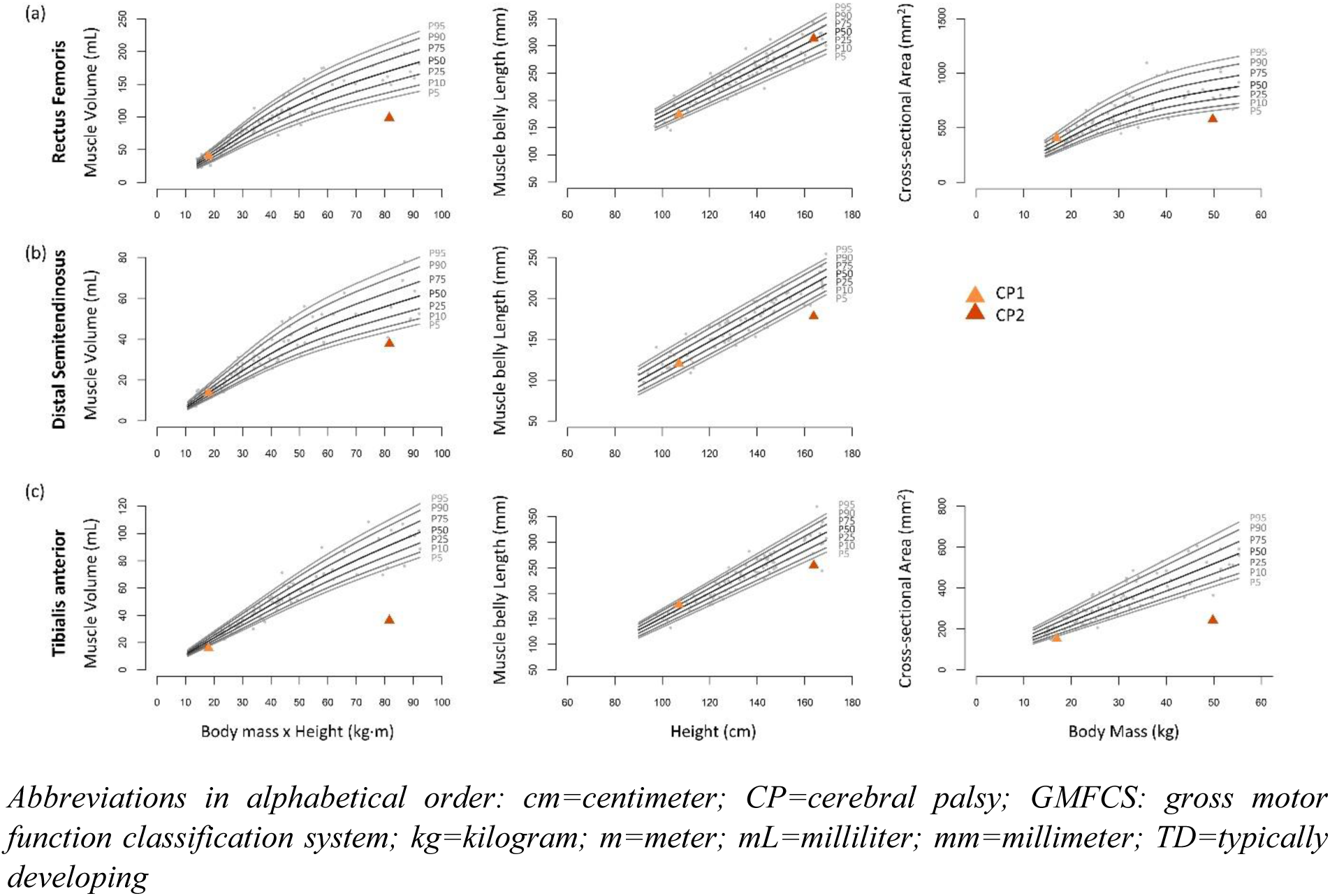
Anthropometric-related TD percentile curves for muscle size of the m. rectus femoris (a), distal m. semitendinosus (b) and m. tibialis anterior (c). Percentiles 5% (P5), 10% (P10), 25% (P25), 50% (P50), 75% (P75), 90% (P90), 95% (P95) were estimated for different values of the product between body mass and height for muscle volume, of height for muscle length and of body mass for cross-sectional area. The observed TD outcomes are plotted in gray. Patient-specific outcomes of two children with CP are visualized by orange triangles and are documented in Table 1. The light orange triangle is the outcome of a patient with GMFCS-level I. The dark orange triangle is the outcome of a patient with GMFCS-level II.

**Table 1:** Absolute muscle size and corresponding z-scores generated from anthropometric-related TD percentile curves in two patients with CP, one classified as GMFCS-level I (CP1) and one classified as GMFCS-level II (CP2)

|  |  |  | **CP1** | **CP2** |
| --- | --- | --- | --- | --- |
| **Age range (years)** |  |  | 5-9 | 10-14 |
| **Sex** |  |  | Female | Female |
| **Body mass (kg)** |  |  | 16.9 | 49.8 |
| **Height (cm)** |  |  | 107.0 | 163.8 |
| **Body mass x Height (kg·m)** |  |  | 18.08 | 81.57 |
| **GMFCS-level** |  |  | I | II |
| **Rectus Femoris** | **Muscle Volume** | **Absolute (mL)** | 40.40 | 98.80 |
|  |  | **z-score** | 0.31 | -2.91 |
|  | **Muscle Length** | **Absolute (mm)** | 174.00 | 313.40 |
|  |  | **z-score** | -0.98 | 0.09 |
|  | **Cross-sectional Area** | **Absolute (mm<sup>2</sup>)** | 406.10 | 578.60 |
|  |  | **z-score** | 0.92 | -2.56 |
| **Distal Semitendinosus** | **Muscle Volume** | **Absolute (mL)** | 13.80 | 37.80 |
|  |  | **z-score** | 0.02 | -2.65 |
|  | **Muscle Length** | **Absolute (mm)** | 120.20 | 178.00 |
|  |  | **z-score** | -0.52 | -3.06 |
| **Tibialis Anterior** | **Muscle Volume** | **Absolute (mL)** | 15.80 | 36.30 |
|  |  | **z-score** | -2.34 | -6.65 |
|  | **Muscle Length** | **Absolute (mm)** | 177.50 | 254.20 |
|  |  | **z-score** | 0.68 | -2.27 |
|  | **Cross-sectional Area** | **Absolute (mm<sup>2</sup>)** | 153.10 | 243.10 |
|  |  | **z-score** | -2.11 | -5.15 |

*Abbreviations in alphabetical order: cm=centimeter; CP=cerebral palsy; GMFCS=gross motor function classification system; kg=kilogram; m=meter; mL=milliliter; mm=millimeter; TD=Typically developing*

### Application 1: Differences in muscle size between two patients with CP

The muscle size assessments of two patients with CP are visualized on the muscle size TD percentile curves in Figure 1 (z-scores in Table 1). CP1 (age between of 5-9 years, female, gross motor function classification system (GMFCS) level I) showed deficits in tibialis anterior volume and cross-sectional area with z-scores below -2, while all other muscle size outcomes were situated on the plotted TD percentile curves with z-scores between -1 and 1. The muscle size outcomes of CP2 (age between 10-14 years, female, GMFCS-level II) were situated below P5 with z-scores below -2, except for the rectus femoris belly length (i.e. z=0.09). Although the absolute muscle size outcomes were larger in CP2 than CP1, CP2 had larger deficits than CP1 in respect to the TD percentile curves (except for the rectus femoris belly length).

### Application 2: Longitudinal follow-up of m. medial gastrocnemius size in a patient with CP

Five m. medial gastrocnemius size assessments of one patient with CP (CP3, age at baseline between 0-4 year, male, GMFCS-level III) during a 2-year follow-up with 6mo intervals are displayed in Figure 2 (z-scores in Table 2). The deficits in volume and cross-sectional area increased during growth, starting above P5 (volume) and P10 (cross-sectional area) to end below P5, while the deficits in muscle length decreased, starting below P5 to end above P5.

**Figure 2:**
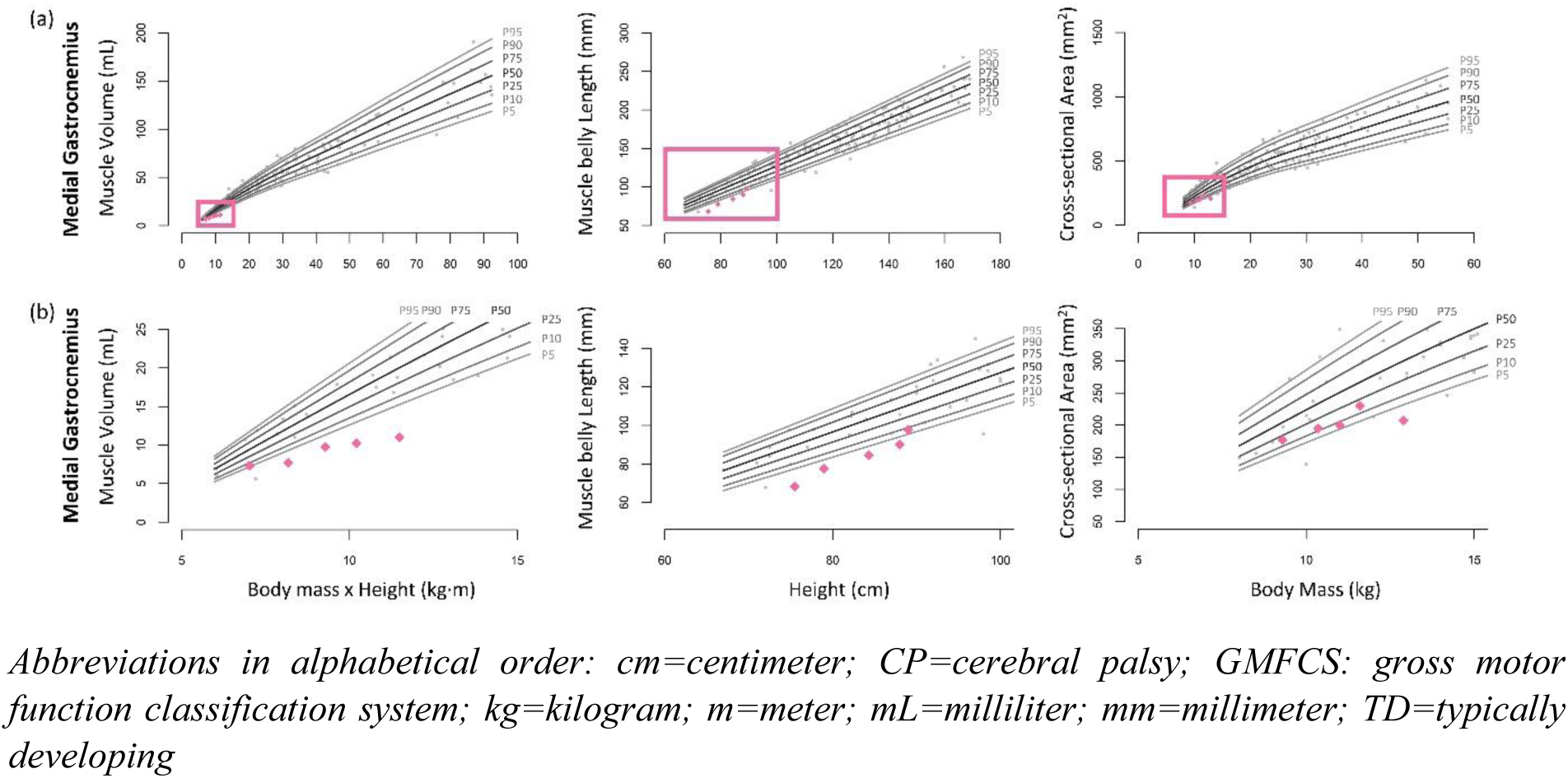
Anthropometric-related TD percentile curves for muscle size of the m. medial gastrocnemius. The full range of the curves are plotted in (a), in (b) we zoomed in on the squares indicated in (a). Percentiles 5% (P5), 10% (P10), 25% (P25), 50% (P50), 75% (P75), 90% (P90), 95% (P95) were estimated for different values of the product between body mass and height for muscle volume, of height for muscle length and of body mass for cross-sectional area. The observed TD outcomes are plotted in gray. Longitudinal patient-specific outcomes of one child with CP (classified as GMFCS-level III) are visualized by pink rotated squares and are documented in Table 2.

**Figure 3:**
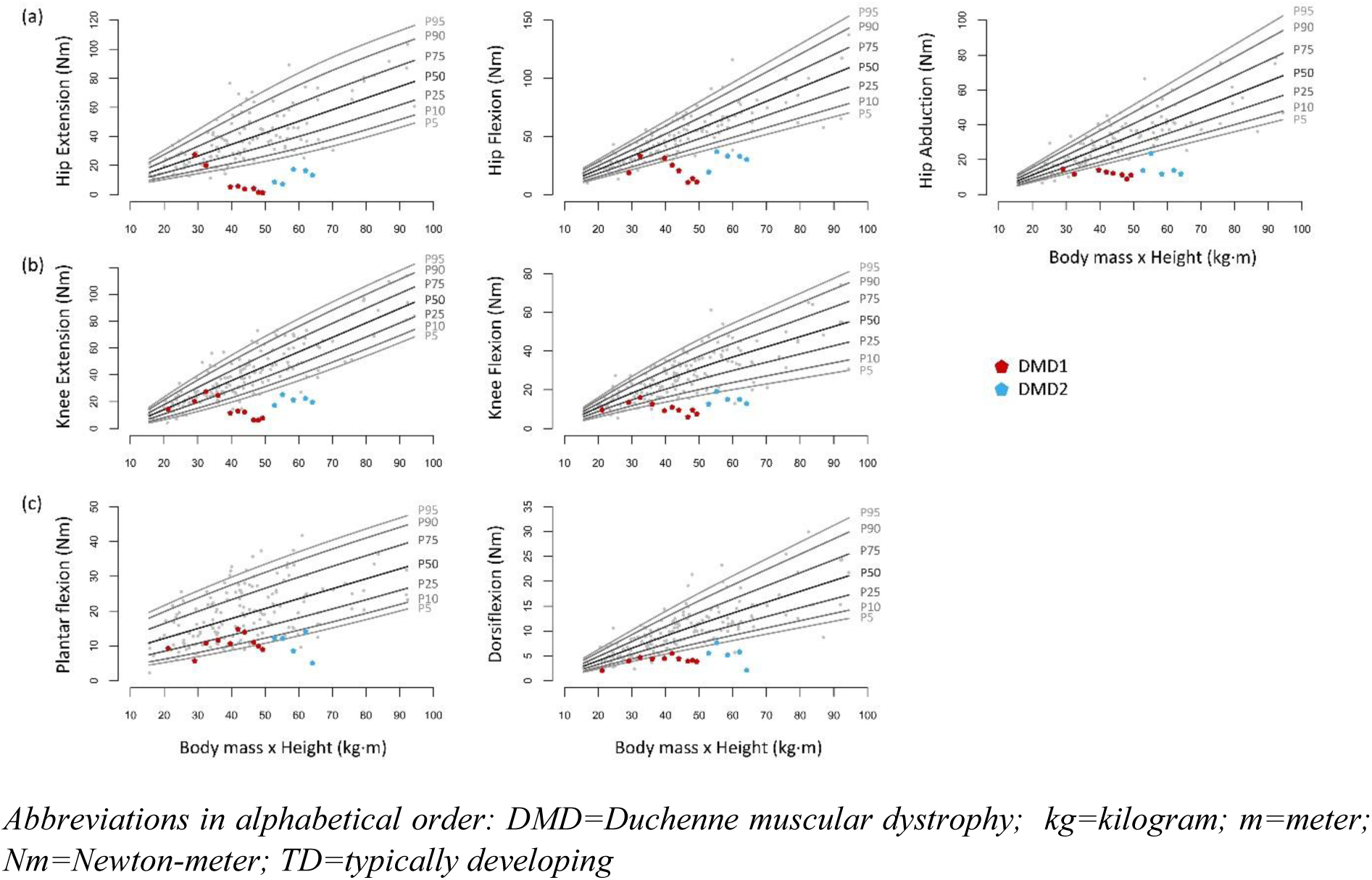
Anthropometric-related TD percentile curves for muscle strength of the hip (a), knee (b) and ankle (c). Percentiles 5% (P5), 10% (P10), 25% (P25), 50% (P50), 75% (P75), 90% (P90), 95% (P95) were estimated for different values of the product between body mass and height. The observed TD outcomes are plotted in gray. Longitudinal patient-specific outcomes of two boys with DMD are visualized by pentagons and are documented in Table 3. The red pentagons are the outcomes of a boy following standard clinical care. The blue pentagons are the outcomes of a boy included in a clinical trial.

**Table 2:** Longitudinal follow-up of absolute muscle size of m. medial gastrocnemius and corresponding z-scores generated from anthropometric-related TD percentile curves in one patient with CP.

|  |  |  | A1 | A2 | A3 | A4 | A5 |
| --- | --- | --- | --- | --- | --- | --- | --- |
| **Age range (years)** |  |  | 0-4 |  |  |  |  |
| **Sex** |  |  | Male |  |  |  |  |
| **Body mass (kg)** |  |  | 9.3 | 10.4 | 11.0 | 11.6 | 12.9 |
| **Height (cm)** |  |  | 75.5 | 79.0 | 84.3 | 88.0 | 89.0 |
| **Body mass x Height (kg·m)** |  |  | 7.02 | 8.18 | 9.27 | 10.21 | 11.48 |
| **GMFCS-level** |  |  | III |  |  |  |  |
| **Medial Gastrocnemius** | **Muscle Volume** | **Absolute (mL)** | 7.34 | 7.74 | 9.74 | 10.26 | 11.02 |
|  |  | **z-score** | -1.54 | -2.48 | -2.33 | -2.66 | -2.96 |
|  | **Muscle Length** | **Absolute (mm)** | 68.45 | 77.60 | 84.48 | 90.19 | 97.74 |
|  |  | **z-score** | -2.78 | -2.20 | -2.17 | -2.07 | -1.41 |
|  | **Cross-sectional Area** | **Absolute (mm<sup>2</sup>)** | 176.89 | 194.75 | 200.36 | 230.23 | 207.30 |
|  |  | **z-score** | -0.93 | -1.16 | -1.44 | -0.95 | -2.33 |

*Abbreviations in alphabetical order: A=assessment; cm=centimeter; CP=cerebral palsy; GMFCS=gross motor function classification system; kg=kilogram; m=meter; mL=milliliter; mm=millimeter; TD=Typically developing*

**Table 3:**
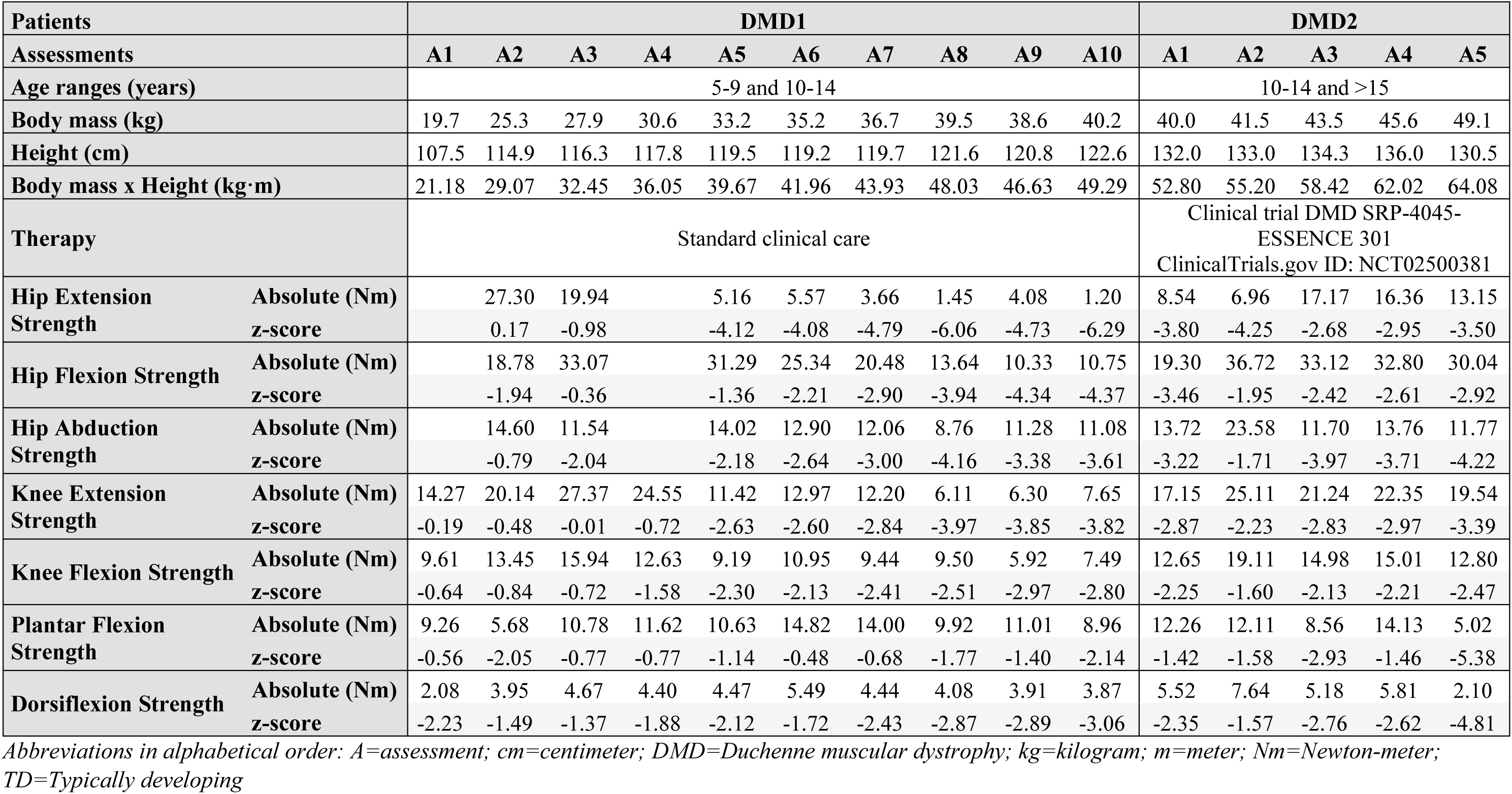
Longitudinal follow-up of absolute muscle strength and corresponding z-scores generated from anthropometric-related TD percentile curves in two boys with DMD, one receiving the standard clinical care (DMD1) and one included in a clinical trial (DMD2)

### Application 3: Longitudinal follow-up of muscle weakness in two patients with DMD

The longitudinal follow-up of muscle weakness in two patients with DMD is visualized on the muscle strength TD percentile curves in Figure 3 (z-scores in Table 3). The first boy (DMD1, age at baseline between 5-9 years) was measured at ten time points during a 6-year follow-up and received standard clinical care. The second boy (DMD2, age at baseline between 10-14 years) was measured at five time points during a 3.5-year follow-up and was included in a clinical trial with exon skipping (DMD SRP-4045-ESSENCE 301, ClinicalTrials.gov ID: NCT02500381). A clear evolution was observed for the strength outcomes of DMD1, from initially being situated on the plotted TD percentile curves to progressively moving away from P5. The z-scores clearly indicated these increasing deficits during growth and showed additional larger deficits for the proximal muscles than the distal muscles in DMD1 (Table 3). Although DMD2 was older at his first assessment than DMD1 at his last assessment, the maximal deficit was smaller in DMD2 and a slower decline was observed comparing the last assessments between the boys in hip and knee extension and flexion strength. Deficits in hip abduction strength were similar between DMD1 and DMD2, while a steep decline in plantar flexion and dorsiflexion strength between the two last assessments was observed in DMD2.

### Application 4: Effect of intervention on muscle strength and volume in a patient with CP

The z-scores for muscle volume and strength outcomes of one patient with CP (CP4, age at baseline between 5-9 years, male, GMFCS-level I) before and after progressive resistance training for the knee extensors, knee flexors and plantar flexors (12 weeks, 3-4 sessions/week, 3 sets of 10 repetitions at 60-80% of the 1-repetition maximum)^29^ are presented in Table 4. The progressive resistance training improved muscle strength, which was indicated by the increase in z-scores. In contrast, the z-scores for muscle volume showed a small decrease, suggesting that the progressive resistance training did not improve muscle volume in CP4.

**Table 4:**
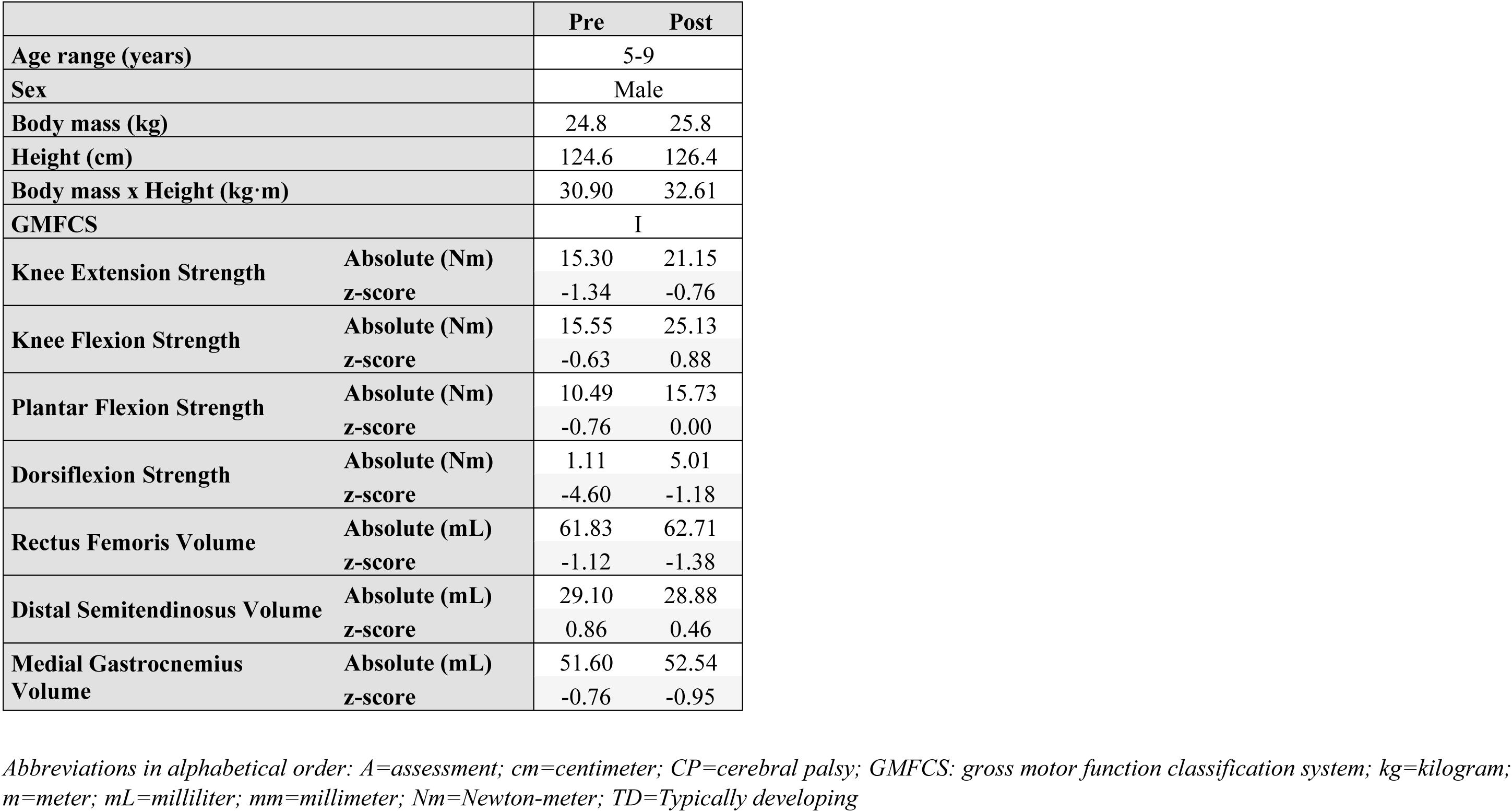
Effect of strength intervention on absolute muscle strength and volume, as well as on the corresponding z-scores generated from anthropometric-related TD percentile curves in one patient with CP.

## Discussion

This study established anthropometric-related percentile curves for muscle size and strength outcomes in a cohort of TD children covering childhood development (0y7mo-17y1mo). The application of these TD percentile curves was demonstrated in growing children with motor disabilities using examples of children with CP and DMD.

The TD percentile curves show a continuous increase of muscle size and strength outcomes with increasing anthropometric parameters. In general, model selection was based on avoiding overfitting and ensuring robustness, while taking biological and clinical expectations of the evolution into account. The continuous increase represents the general maturation of the musculoskeletal system, which was expected since muscle parameters increase substantially during childhood to follow the growth of skeletal bones and to generate the power required to move the growing body.^1,2^ Previous studies often presented the relationship between muscle size and strength outcomes as a linear relationship with age or anthropometric growth, and therefore, these outcomes were often “linearly” normalized to anthropometric growth.^25^ However, the present curvilinear outcomes, specifically for muscle volume and cross-sectional area, indicate the errors and misinterpretations that can be made when assuming a purely linear relationship between these parameters.

Small differences in the fitted models between outcomes were visible. While muscle length is mostly governed by skeletal growth, muscle volume and cross-sectional area are also impacted by hormonal status, nutrition, physical activity and sports participation, inducing more variability^30^. Since strength is related to muscle size, this also explains the high variability in muscle strength^3^. Additionally, muscle strength evaluations require a certain level of understanding and cooperation, as well as neural control of muscle activation, causing already a higher variability in young children.

The included examples of children with CP and DMD demonstrate the clinical applications of the presented anthropometric-related TD percentile curves. These curves allow us to express patient-specific outcomes into deficits (z-scores) in reference to TD peers. Thereby, normal muscle growth and strength development are accounted for, resulting in the sole depiction of pathological alterations. The use of these deficits, namely the non-dimensional z-scores, facilitates the comparison of pathology severity between patients, as well as the delineation of pathological trajectories during growth within and across patients. Therefore, these deficits could be valuable in clinical research as well as for patient-specific follow-up in clinical practice. Within a patient, the non-dimensional z-scores allows for the comparison of pathological involvement between muscles and muscle groups, for example the proximal vs. the distal involvement in boys with DMD. In addition, the deficits between different muscle size parameters can be compared, aiding in the understanding of the contribution of altered longitudinal and cross-sectional dimensions to altered muscle growth. Moreover, combining the size and strength outcomes can contribute to the determination of the origin of muscle weakness in children with CP, i.e., mostly muscle size-related or more impacted by neural control^3^. Lastly, interpreting the impact of treatments could be improved by using the TD percentile curves to separate the treatment effect from the effect of growth itself. Consequently, these deficits may constitute of sensitive outcome measures to prove, for example, the efficacy of clinical trials aiming at slowing down disease progression in boys with DMD. In the same line, these deficits could be used to monitor the effect of treatment included in the standard clinical care, such as strength and the tone-reducing interventions in children with CP, and potentially improving patient-specific clinical decision-making in different pediatric pathologies.

In the future, pathology-specific percentile curves would allow the comparison of a child with CP or DMD with peers with CP or DMD, respectively. However, in the absence of large pathology-specific databases, this comparison between patients with the same pathology can be performed through z-scores based on the current TD percentile curves. These pathology-specific percentile curves could be complementary to the current TD percentile curves, the latter showing the extent of muscular pathology in reference to a TD population and the former indicating the position of the child in reference to a peer group. The future application of these percentile curves on larger pathology-specific databases might unravel the presentation of atypical muscle strength and size development, providing biomarkers for earlier disease detection and for increased understanding of the link with altered motor function.

Although the clinical applications are multifaceted, this investigation has some limitations. Similar to previous percentile curves^20^, the proposed TD percentile curves were based solely on cross-sectional data. Incorporating longitudinal data in the future will strengthen descriptions of growth, especially for pathology-specific databases. Moreover, the number of children was not equally distributed across anthropometric values. In children with very low anthropometric values, only the muscle size parameters of the medial gastrocnemius were available, and the application of 3DfUS to other muscles must be explored in these children. Moreover, strength assessments were only possible from the age of 4-5 years onwards, therefore, these TD percentile curves start from higher anthropometric values. Overall, the number of participants with higher anthropometric values is limited, hence these end-parts of the TD percentile curves should be interpreted with caution. Finally, the number of children per sex subgroup was too small to create percentile curves, therefore male and female participants were combined in one dataset. Future studies should define the influence of sex and onset of puberty on the evolution of muscle size and strength with growth.

We have established anthropometric-related percentile curves for muscle size and strength in a cohort of TD children covering childhood development. The clinical applications show the ability to track the trajectory of muscle alterations and weakness in children with CP and boys with DMD and allow for the evaluation of the impact of treatment on muscle size and function, while taking into account the expected typical change due to natural growth. The online graphical user interface (https://cmal.shinyapps.io/Z-score_calculator_MG_muscle_volume/) facilitates the use of the established TD percentile curves across multiple centers, allowing multi-center studies and discussions.

## Data Availability

All data produced in the present study are available upon reasonable request to the authors

## List of abbreviations

CP: cerebral palsy
DMD: Duchenne muscular dystrophy
GAMLSS: generalized additive models for location, scale and shape
TD: typically developing

## Supporting information

The following additional material may be found online.

**Appendix S1**: Details on the distribution parameters of the generalized additive models for location, scale and shape for muscle size outcomes

**Appendix S2**: Details on the distribution parameters of the generalized additive models for location, scale and shape for muscle strength outcomes

**Online graphical user interface**: https://cmal.shinyapps.io/Z-score_calculator_MG_muscle_volume/

## Acknowledgements

The authors wish to thank all the children and parents for their participation in this study and the institutional boards of the school for their permission to perform measurements at their school. We also thank the colleagues of the University Hospitals of Leuven and all graduate students from the KU Leuven and the University of Ghent involved in the recruitment and data collection. A special thank you to Ester Huyghe and Elze Stoop for their support in data collection and processing.

## Reference list

1. Peeters N, Hanssen B, De Beukelaer N, et al. A comprehensive normative reference database of muscle morphology in typically developing children aged 3–18 years—a cross-sectional ultrasound study. J Anat 2023; 242: 754–70.

2. Beunen G, Thomis M. Muscular Strength Development in Children and Adolescents. Human Kinetics Publishers, Inc, 2000.

3. Hanssen B, Peeters N, Vandekerckhove I, et al. The Contribution of Decreased Muscle Size to Muscle Weakness in Children With Spastic Cerebral Palsy. Front Neurol 2021; 12. DOI:10.3389/fneur.2021.692582.

4. De Beukelaer N, Vandekerckhove I, Molenberghs G, et al. Longitudinal trajectory of medial gastrocnemius muscle growth in the first years of life. Dev Med Child Neurol 2023. DOI:10.1111/dmcn.15763.

5. Verreydt I, Vandekerckhove I, Stoop E, et al. Instrumented strength assessment in typically developing children and children with a neural or neuromuscular disorder: A reliability, validity and responsiveness study. Front Physiol 2022; 13. DOI:10.3389/fphys.2022.855222.

6. De Beukelaer N, Vandekerckhove I, Huyghe E, et al. Morphological Medial Gastrocnemius Muscle Growth in Ambulant Children with Spastic Cerebral Palsy: A Prospective Longitudinal Study. J Clin Med 2023; 12: 1564.

7. Sussman M. Duchenne Muscular Dystrophy. J Am Acad Orthop Surg 2002; 10: 138–51.

8. Surveillance of cerebral palsy in Europe: a collaboration of cerebral palsy surveys and registers. Dev Med Child Neurol 2000; 42: 816–24.

9. McIntyre S, Goldsmith S, Webb A, et al. Global prevalence of cerebral palsy: A systematic analysis. Dev Med Child Neurol 2022; 64: 1494–506.

10. Graham HK, Rosenbaum P, Paneth N, et al. Cerebral palsy. Nat Rev Dis Primers 2016; 2: 15082.

11. Handsfield GG, Williams S, Khuu S, Lichtwark G, Stott NS. Muscle architecture, growth, and biological Remodelling in cerebral palsy: a narrative review. BMC Musculoskelet Disord 2022; 23: 1–17.

12. Mockford M, Caulton JM. The Pathophysiological Basis of Weakness in Children with Cerebral Palsy. Pediatric Physical Therapy 2010; 22: 222–33.

13. Bushby K, Finkel R, Birnkrant DJ, et al. Diagnosis and management of Duchenne muscular dystrophy, part 1: diagnosis, and pharmacological and psychosocial management. Lancet Neurol 2010; 9: 77–93.

14. Sutherland D. PATHOMECHANICS OF GAIT IN DUCHENNE MUSCULAR DYSTROPHY. Journal of Pediatric Orthopaedics 1981; 1. DOI:10.1097/01241398-198111000-00055.

15. Goudriaan M, Nieuwenhuys A, Schless SH, Goemans N, Molenaers G, Desloovere K. A new strength assessment to evaluate the association between muscle weakness and gait pathology in children with cerebral palsy. In: PLoS ONE. Public Library of Science, 2018: e0191097.

16. Cenni F, Monari D, Desloovere K, Aertbeliën E, Schless SH, Bruyninckx H. The reliability and validity of a clinical 3D freehand ultrasound system. Comput Methods Programs Biomed 2016; 136. DOI:10.1016/j.cmpb.2016.09.001.

17. Hanssen B, Peeters N, Dewit T, et al. Reliability of 3D freehand ultrasound to assess lower limb muscles in children with spastic cerebral palsy and typical development. J Anat 2023. DOI:10.1111/joa.13839.

18. Stimpson G, Raquq S, Chesshyre M, et al. Growth pattern trajectories in boys with Duchenne muscular dystrophy. Orphanet J Rare Dis 2022; 17: 1–11.

19. Day SM, Strauss DJ, Vachon PJ, Rosenbloom L, Shavelle RM, Wu YW. Growth patterns in a population of children and adolescents with cerebral palsy. Dev Med Child Neurol 2007; 49: 167–71.

20. Borghi E, de Onis M, Garza C, et al. Construction of the World Health Organization child growth standards: selection of methods for attained growth curves. Stat Med 2006; 25: 247–65.

21. Cenni F, Monari D, Desloovere K, Aertbeliën E, Schless SH, Bruyninckx H. The reliability and validity of a clinical 3D freehand ultrasound system. Comput Methods Programs Biomed 2016; 136: 179–87.

22. Cenni F, Schless SH, Monari D, et al. An innovative solution to reduce muscle deformation during ultrasonography data collection. J Biomech 2018; 77: 194–200.

23. Stasinopoulos DM, Rigby RA. Generalized additive models for location scale and shape (GAMLSS) in R. J Stat Softw 2007; 23: 1–46.

24. Rigby RA, Stasinopoulos DM. Generalized additive models for location, scale and shape. J R Stat Soc Ser C Appl Stat 2005; 54: 507–54.

25. Williams SA, Stott NS, Valentine J, Elliott C, Reid SL. Measuring skeletal muscle morphology and architecture with imaging modalities in children with cerebral palsy: a scoping review. Dev Med Child Neurol 2021; 63: 263–73.

26. Cole TJ, Green PJ. Smoothing reference centile curves: the LMS method and penalized likelihood. Stat Med 1992; 11: 1305–19.

27. Rigby RA, Stasinopoulos DM. Smooth centile curves for skew and kurtotic data modelled using the Box–Cox power exponential distribution. Stat Med 2004; 23: 3053–76.

28. Rigby RA, Stasinopoulos DM. Using the Box-Cox t distribution in GAMLSS to model skewness and kurtosis. Stat Modelling 2006; 6: 209–29.

29. Hanssen B, Peeters N, De Beukelaer N, et al. Progressive resistance training for children with cerebral palsy: A randomized controlled trial evaluating the effects on muscle strength and morphology. Front Physiol 2022; 13. DOI:10.3389/fphys.2022.911162.

30. Sartori R, Romanello V, Sandri M. Mechanisms of muscle atrophy and hypertrophy: implications in health and disease. DOI:10.1038/s41467-020-20123-1.

