## Appendix S1 for "Anthropometric-related percentile curves for muscle size and strength of lower limb muscles of typically developing children"

|  |  |  |  |  |  | **Model** | | | | | |
| --- | --- | --- | --- | --- | --- | --- | --- | --- | --- | --- | --- |
|  |  | **n subj** | **n ♂/♀** | **Age Range** | **Anthropometrics Range** | **Distr** |  | **Parameters** | | | |
| **Outcomes** | |  |  |  |  |  |  | **µ** | **log(σ)** | **ν** | **log(τ)** |
| Rectus Femoris | Muscle Volume (mL) | 79 | 36/43 | 3:0 – 17:10  y:mo | 14.1 - 92.1  kg·m | BCCG |  | pb(Body Mass·Height) | ~1 | ~1 |  |
|  |  |  |  |  |  |  |  | *df= 1.89* | *-1.89* | *0.81* |  |
|  | Muscle Length (mm) | 79 | 36/43 | 3:0 – 17:10  y:mo | 97 - 169  cm | BCCG |  | Height | ~1 | ~1 |  |
|  |  |  |  |  |  |  |  | *2.19**** | *-2.64* | *1.10* |  |
|  | Cross-sectional Area (mm^2^) | 77 | 35/42 | 3:0 – 17:10  y:mo | 14.5 - 55.3  kg | BCCG |  | pb(Body Mass) | ~1 | ~1 |  |
|  |  |  |  |  |  |  |  | *df= 1.60* | *-1.84* | *-0.38* |  |
| Distal Semitendinosus | Muscle Volume (mL) | 90 | 47/43 | 2:1 – 17:10 y:mo | 10.8 - 92.1  kg·m | BCCG |  | pb(Body Mass·Height) | ~1 | ~1 |  |
|  |  |  |  |  |  |  |  | *df= 1.79* | *-1.84* | *-0.24* |  |
|  | Muscle Length (mm) | 92 | 48/44 | 2:1 – 17:10 y:mo | 90 - 169  cm | BCCG |  | Height | Height | ~1 |  |
|  |  |  |  |  |  |  |  | *1.61**** | *-0.01** | *0.64* |  |
| Tibialis Anterior | Muscle Volume (mL) | 84 | 40/44 | 3:0 – 16:1 y:mo | 10.8 - 92.4  kg·m | BCCG |  | pb(Body Mass·Height) | ~1 | ~1 |  |
|  |  |  |  |  |  |  |  | *df= 1.39* | *-2.14* | *0.36* |  |
|  | Muscle Length (mm) | 84 | 40/44 | 3:0 – 16:1 y:mo | 90 - 169  cm | BCCG |  | Height | ~1 | ~1 |  |
|  |  |  |  |  |  |  |  | *2.42**** | *-2.64* | *1.59* |  |
|  | Cross-sectional Area (mm^2^) | 84 | 40/44 | 3:0 – 16:1 y:mo | 12.0 - 55.3  kg | BCCG |  | Body Mass | ~1 | ~1 |  |
|  |  |  |  |  |  |  |  | *9.40**** | *-1.93* | *0.02* |  |
| Medial Gastrocnemius | Muscle Volume (mL) | 142 | 77/65 | 0:7 – 16:1 y:mo | 6.0 - 92.4  kg·m | BCT |  | pb(Body Mass·Height) | ~1 | ~1 | ~1 |
|  |  |  |  |  |  |  |  | *df= 2.58* | *-1.95* | *0.89* | *3.24* |
|  | Muscle Length (mm) | 145 | 81/64 | 0:7 – 16:1 y:mo | 67 - 169  cm | BCCG |  | Height | ~1 | ~1 |  |
|  |  |  |  |  |  |  |  | *1.54**** | *-2.54* | *1.61* |  |
|  | Cross-sectional Area (mm^2^) | 143 | 78/65 | 0:7 – 16:1 y:mo | 8.0 - 55.3  kg | BCCG |  | pb(Body Mass) | ~1 | ~1 |  |
|  |  |  |  |  |  |  |  | *df= 2.44* | *-1.88* | *0.21* |  |

**Appendix S1:** Details on the distribution parameters of the generalized additive models for location, scale and shape for muscle size outcomes

*p<0.05; **p<0.01; ***p<0.0001

The modelled median (µ), coefficient of variation (σ), skewness (ν) and kurtosis (τ) of the Box-Cox Cole Green (BCCG) or Box-Cox t (BCT) distribution of the muscle size outcomes with respect to anthropometrics are documented. The model’s formula is provided in the upper row and the estimates or additional degrees of freedom in the lower row (*in italic*) per muscle size outcome. Body mass*height is the explanatory variable for muscle volume, height for muscle length and body mass for cross-sectional area. The effect of anthropometrics is modelled either as a P-spline (i.e., pb()), as a linear function (i.e., body mass*height, height or body mass) or as a constant (~1). The additional degrees of freedom (df) are given in case of a P-spline, the regression coefficient in case of a linear function and the intercept in case of a constant. The number of subjects, ratio boys/girls, age range and anthropometric values range are described per model. The number of subjects differed between outcomes as a result of available data, cut-off based on anthropometric values and outliers.

BCCG=Box-Cox Cole Green; BCT=Box-Cox t; cm=centimeter; df=degrees of freedom; Distr=distribution; kg=kilogram; log()=logarithm; m=meter; mL=milliliter; mm=millimeter; mo=months; n=number; pb()=P-spline; subj=subjects; y=years; µ=median; σ=coefficient of variation; ν=skewness; τ=kurtosis
