## Appendix S2 for "Anthropometric-related percentile curves for muscle size and strength of lower limb muscles of typically developing children"

**Appendix S2**: Details on the distribution parameters of the generalized additive models for location, scale and shape for muscle strength outcomes

|  |  |  |  |  |  | **Model** | | | | | |
| --- | --- | --- | --- | --- | --- | --- | --- | --- | --- | --- | --- |
|  |  | **n subj** | **n ♂/♀** | **Age Range** | **Anthropometrics Range** | **Distr** |  | **Parameters** | | | |
| **Outcomes** | |  |  |  |  |  |  | **µ** | **log(σ)** | **ν** | **log(τ)** |
| Hip | Extension Strength (Nm) | 124 | 90/34 | 4.5 - 16.1 years | 15.6 - 94.4  kg·m | BCCG |  | Body Mass·Height | pb(Body Mass·Height) | ~1 |  |
|  |  |  |  |  |  |  |  | *0.80**** | *df=0.69* | *0.30* |  |
|  | Flexion Strength (Nm) | 123 | 89/34 | 4.5 - 16.1 years | 15.6 - 94.4  kg·m | BCCG |  | Body Mass·Height | ~1 | ~1 |  |
|  |  |  |  |  |  |  |  | *1.18**** | *-1.46* | *0.64* |  |
|  | Abduction Strength (Nm) | 124 | 90/34 | 4.5 - 16.1 years | 15.6 - 94.4  kg·m | BCCG |  | Body Mass·Height | ~1 | ~1 |  |
|  |  |  |  |  |  |  |  | *0.77**** | *-1.33* | *0.29* |  |
| Knee | Extension Strength (Nm) | 153 | 108/45 | 4.5 - 16.1 years | 15.6 - 94.4  kg·m | BCCG |  | Body Mass·Height | Body Mass·Height | ~1 |  |
|  |  |  |  |  |  |  |  | *1.08**** | *-0.01*** | *0.66* |  |
|  | Flexion Strength (Nm) | 153 | 108/45 | 4.5 - 16.1 years | 15.6 - 94.4  kg·m | BCCG |  | pb(Body Mass·Height) | ~1 | ~1 |  |
|  |  |  |  |  |  |  |  | *df=1.19* | *-1.26* | *0.93* |  |
| Ankle | Plantar Flexion Strength (Nm) | 150 | 107/43 | 4.5 - 16.1 years | 15.6 - 92.4  kg·m | BCPE |  | Body Mass·Height | Body Mass·Height | ~1 | ~1 |
|  |  |  |  |  |  |  |  | *0.29**** | *-0.01** | *0.55* | *1.31* |
|  | Dorsiflexion Strength (Nm) | 151 | 106/45 | 4.5 - 16.1 years | 15.6 - 94.4  kg·m | BCCG |  | pb(Body Mass·Height) | ~1 | ~1 |  |
|  |  |  |  |  |  |  |  | *df=0.56* | *-1.24* | *0.36* |  |

*p<0.05; **p<0.01; ***p<0.0001

The modelled median (µ), coefficient of variation (σ), skewness (ν) and kurtosis (τ) of the Box-Cox Cole Green (BCCG) or Box-Cox power exponential (BCPE) distribution of the muscle strength outcomes with respect to anthropometrics are documented. The model’s formula is provided in the upper row and the estimates or additional degrees of freedom in the lower row (*in italic*) per muscle strength outcome. Body mass*height is the explanatory variable for muscle strength. The effect of anthropometrics is modelled either as a P-spline (i.e., pb()), as a linear function (i.e., body mass*height) or as a constant (~1). The additional degrees of freedom (df) are given in case of a P-spline, the regression coefficient in case of a linear function and the intercept in case of a constant. The number of subjects, ratio boys/girls, age range and anthropometric values range are described per model. The number of subjects differed between outcomes as a result of available data, cut-off based on anthropometric values and outliers.

BCCG, Box-Cox Cole Green; BCPE, Box-Cox power exponential; df, degrees of freedom; Distr, distribution; kg, kilogram; log(), logarithm; m, meter; n, number; pb(), P-spline; subj, subjects; µ, median; σ, coefficient of variation; ν, skewness; τ, kurtosis
